# Identification of Acute Respiratory Distress Syndrome subphenotypes denovo using routine clinical data: a retrospective analysis of ARDS clinical trials

**DOI:** 10.1101/2021.07.08.21260102

**Authors:** Abhijit Duggal, Rachel Kast, Emily Van Ark, Lucas Bulgarelli, Matthew T. Siuba, Jeff Osborn, Diego Rey, Fernando G Zampieri, Alexandre B Cavalcanti, Israel S Maia, Denise M Paisani, Ligia N Laranjeira, Ary Serpa Neto, Rodrigo Octávio Deliberato

**Affiliations:** Department of Critical Care Medicine, Respiratory Institute, Cleveland Clinic, Cleveland, Ohio, USA; Department of Clinical Data Science, Endpoint Health Inc, Palo Alto, California, USA; HCor Research Institute, São Paulo, Brazil; Australian and New Zealand Intensive Care Research Centre (ANZIC-RC), School of Public Health and Preventive Medicine, Monash University, Melbourne, Australia; Department of Critical Care, Melbourne Medical School, University of Melbourne, Austin Hospital, Melbourne, Australia; Department of Intensive Care, Austin Hospital, Melbourne, Australia; Data Analytics Research and Evaluation (DARE) Centre, Austin Hospital, Melbourne, Australia; Department of Critical Care Medicine, Hospital Israelita Albert Einstein, São Paulo, Brazil

**Author notes:** **Correspondence:** Abhijit Duggal MD, Address: 9500 Euclid Ave, L2-330, Cleveland, Ohio, 44195. Authors contributed equally (Abhijit Duggal, Rachel Kast, Emily Van Ark, Lucas Bulgarelli, Rodrigo Octávio Deliberato).

**Keywords:** Subphenotype, machine learning, ARDS, critical care, clinical data, clustering

## Abstract

**Rationale:** The acute respiratory distress syndrome (ARDS) is a heterogenous condition, and identification of subphenotypes may help in better risk stratification.

**Objectives:** Identify ARDS subphenotypes using new simpler methodology and readily available clinical variables.

**Design:** Retrospective Cohort Study of ARDS trials.

**Setting:** Data from the U.S. ARDSNet trials and from the international ART trial.

**Participants:** 3763 patients from ARDSNet datasets and 1010 patients from the ART dataset.

**Primary and secondary outcome measures:** The primary outcome was 60-day or 28-day mortality, depending on what was reported in the original trial. K-means cluster analysis was performed to identify subgroups. For feature selection, sets. Sets of candidate variables were tested to assess their ability to produce different probabilities for mortality in each cluster. Clusters were compared to biomarker data, allowing identification of subphenotypes.

**Results:** Data from 4,773 patients was analyzed. Two subphenotypes (A and B) resulted in optimal separation in the final model, which included nine routinely collected clinical variables, namely: heart rate, mean arterial pressure, respiratory rate, bilirubin, bicarbonate, creatinine, PaO_2_, arterial pH, and FiO_2_. Participants in subphenotype B showed increased levels of pro-inflammatory markers, had consistently higher mortality, lower number of ventilator-free days at day 28, and longer duration of ventilation compared to patients in the subphenotype A.

**Conclusions:** Routinely available clinical data can successfully identify two distinct subphenotypes in adult ARDS patients. This work may facilitate implementation of precision therapy in ARDS clinical trials.

**ARTICLE SUMMARY:** *Strengths and limitations of this study:*

- Largest cohort of patients used to identify subphenotypes of ARDS patients.
- Subphenotypes were validated in the population of a large international ARDS randomized controlled trial.
- Subphenotypes were identified by using only routinely collected clinical data.
- Our use of data exclusively from randomized controlled trials does not prove generalizability to unselected ARDS populations.
- The clinical utility of the subphenotypes have to be validated in a prospective study.

## INTRODUCTION

The Berlin definition of acute respiratory distress syndrome (ARDS) encompasses acute hypoxemic respiratory failure due to a wide variety of etiologies [1]. Due to this inclusion of heterogeneous conditions within the syndrome, there are significant clinical and biological differences that makes ARDS challenging to treat [2,3]. These differences amongst ARDS patients are associated with variation in risk of disease development and progression [3,4], potentially generating differential responses to treatments and interventions [5–10]. In spite of those evidences, clinical risk stratification of ARDS patients still solely depends on PaO_2_/FiO_2_ ratios [11,12], possibly misleading the interpretation of results in clinical trials and clinicians when evaluating treatment options for patients [13].

Therefore, identifying groups of patients who have similar clinical, physiologic, or biomarker traits becomes relevant [6,14] as it can help with stratification of patients producing better targeted therapies and interventions [15]. These different groups can be defined as ARDS subphenotypes [4,14]. Two ARDS subphenotypes have been consistently identified in previous studies [6–10,16–18]. However, these models are complex, and significant barriers exist in their implementation and use in clinical practice. Existing models use up to 40 predictor variables, including biomarkers and other variables that are not readily available at the bedside [6–10,16–18]. These limitations explain the current status quo of ARDS care, where clinicians must depend on the limited prognostic value of PaO_2_/FiO_2_ ratios instead of biologically distinct subphenotypes.

We hypothesized that the use of a simpler methodology and a small number of easily available clinical variables could identify new ARDS subphenotypes and thus provide the means to allow future implementation of bedside stratification.

## METHODS

### Data source and participants

We performed a retrospective study using a de-identified dataset pooling data from six randomized clinical trials in patients with ARDS, namely: ARMA, ALVEOLI, FACTT, EDEN, SAILS, and ART [19–24]. Patients in ARMA, ALVEOLI, FACTT, EDEN and SAILS trials were eligible if they met the American-European consensus for ARDS, including patients with a PaO_2_ / FiO_2_ ratio < 300 up to 48 hours before enrollment. From 1996 to 2013, these trials enrolled 902, 549, 1000, 1000, and 745 patients, respectively, and tested a variety of interventions [19–23]. Between 2011 and 2017 the international ART study enrolled 1010 adult patients diagnosed with moderate to severe ARDS according to the Berlin definition (PaO_2_ / FiO_2_ ratio < 200) for less than 72 hours of duration and assessed two different ventilatory strategies [24]. To avoid biases due to high mortality in the high tidal volume group of the ARMA study [19], which has not been standard of care since the beginning of 2000, only 473 patients receiving low tidal volume in that study were included.

### Predictors

Six clinical trials were assessed to identify a set of clinical variables recorded closest to time of randomization which were most commonly available across all datasets. The list of potential candidates was then further refined to include only those that are frequently observed in the routine care of ARDS patients at the time of its diagnosis. In order to develop a clustering algorithm for potential rapid translation into clinical use, elements which would not be commonly found in the electronic health records (EHR) at the time of ARDS diagnosis, such as biomarker levels, ARDS risk factors, organ support apart from mechanical ventilation settings, and severity scores, were excluded from model development. The treatment assignment in the original trials, and clinical outcomes were not considered in the model development.

After all assessment, 16 variables that are routinely collected as part of the usual care and which were uniformly present in all the trials were considered, including: age, gender, arterial pH, PaO_2_, PaCO_2_, bicarbonate, creatinine, bilirubin, platelets, heart rate, respiratory rate, mean arterial pressure, positive end-expiratory pressure (PEEP), plateau pressure, FiO_2_, and tidal volume adjusted for predicted body weight (mL/kg PBW). The PBW was calculated as equal to 50 + 0.91 (centimeters of height – 152.4) in males, and 45.5 + 0.91 (centimeters of height – 152.4) in females [18]. These variables were grouped into five domains named demographics, arterial blood gases, laboratory values, vital signs, and ventilatory variables. Plateau pressure was excluded due to a high rate of missingness across the trials included in the training set. Amount of missing data in the training datasets is reported in **eTable 1**.

### Outcomes

The primary outcome was 60-day mortality for all ARDSnet trials, and 28-day mortality for ART trial. Secondary outcomes included 90-day mortality, number of ventilator free days at day 28 [25], and the duration of mechanical ventilation in survivors within the first 28 days post enrollment.

### Data preparation

Data preprocessing was performed before modeling, and the pooled dataset was assessed for completeness and consistency. Patients with values out of the plausible physiological range for a specific variable were excluded from the final analysis (described in **eTable 2**). The training dataset was constructed using data from the two largest ARDSnet trials, EDEN and FACTT. The validation dataset was sourced from the four remaining trials: ALVEOLI, ARMA, SAILS, and ART. Means and standard deviations for z-scoring variables were calculated from the training dataset and subsequently applied to the validation data.

### Statistical analysis

Baseline and outcome data were presented according to the assigned cluster. Continuous variables were presented as medians with their interquartile ranges and categorical variables as total number and percentage. Proportions were compared using Fisher exact tests and continuous variables were compared using the Wilcoxon rank-sum test. Study outcomes were further compared using the median and mean absolute differences for continuous and categorical values, respectively.

### Model development and validation

For the model development, the K-means clustering algorithm was used. K-means is one of the simplest and most commonly used classes of clustering algorithms. In critical care research, unsupervised machine learning techniques have already been used in several studies, attempting to find homogeneous subgroups within a broad heterogeneous population [26]. This specific algorithm identifies a K number of clusters in a dataset by finding K centroids within the n-dimensional space of clinical features [26].

For feature selection, different sets of candidate variables were tested to assess their ability to produce significantly different mortality probabilities in each cluster using the minimum amount of readily available clinical data. For each set of candidate variables, the optimal number of clusters was determined by comparing models with between 2 and 5 clusters, using the Elbow method [27] and the Calinski-Harabasz index [28]. Information about the methods for selecting number of clusters are provided in the supplemental material.

The following steps were performed for the final model selection: 1) all predictors were assessed for correlation (**eTable 3**); and 2) ten different combinations of the proposed variables were investigated. These combinations were developed based on the perceived clinical importance of each variable and its combinations. All 10 models were tested for the optimal number of clusters and based on both the Elbow method and the Calinski-Harabasz index, as described above. The models were then compared, aiming for the minimum set of variables with high 60-day mortality separation. The description of each model is show in **eTable 4**.

Biological and clinical characteristics of the clusters were evaluated using clinical, laboratory, and (when available) biomarker data to establish subphenotypes [4]. All iterations in model development were done on the training set and the generalizability of the final model was assessed using the validation dataset. K-means clustering analysis is structured to ignore cases with missing data. No assumption was made for missingness and we therefore conducted a complete case analysis. Model development and evaluation was performed using Python version 3.8 and scikit-learn 0.23.1.

### Data availability

Data from the ARDSnet studies (EDEN, FACTT, ARMA, ALVEOLI and SAILS) is publicly available from the NHLBI ARDS Network and data from the ART trial can be requested from study authors.

## RESULTS

### Participants

Data from 4777 clinical trial patients were considered for inclusion. In total, 4 patients were excluded for having clinical measurements outside plausible range. The remaining 1998 patients from EDEN and FACTT trials were included in the training set, while the 2775 patients from ARMA, ALVEOLI, SAILS, and ART were included in the validation cohort.

Baseline characteristics of the patients in the training and validation sets are presented in **Table 1**. Pneumonia was the prevailing etiology followed by sepsis and aspiration in all trials. Between 29.3% to 72.7% of the patients were receiving vasopressors at the time of randomization. At randomization, PaO_2_ / FiO_2_ ratio ranged from 112 (75 - 158) to 134 (96 - 185) mmHg, and PEEP from 8 (5 - 10) to 12 (10 - 14) cmH_2_O across trials. Mortality at 60 days for the ARDSnet trials ranged from 22.7% to 30.1%, while in the ART trial mortality at 28 days was 58.8%.

**Table 1.** Baseline Characteristics and Clinical Outcomes in the Included Trials.

|  |  |  |  |  |  |  |
| --- | --- | --- | --- | --- | --- | --- |
|  | Training set (n = 1998) |  | Validation set (n = 2775) |  |  |  |
|  | EDEN<br>(n = 1000) | FACTT<br>(n = 998) | ALVEOLI<br>(n = 549) | ARMA<br>(n = 472) | ART<br>(n = 1010) | SAILS<br>(n = 744) |
| Age, year | 52.0 (42.0 - 63.0) | 49.0 (38.0 - 60.8) | 50.0 (39.0 - 65.0) | 50.0 (37.8 - 65.0) | 52.0 (36.0 - 64.0) | 55.0 (42.0 - 66.0) |
| Male gender - no. (%) | 510 (51.0) | 533 (53.4) | 302 (55.0) | 285 (60.4) | 631 (62.5) | 365 (49.0) |
| Etiology - no. (%) |  |  |  |  |  |  |
| Pneumonia | 650 (65.0) | 471 (47.2) | 221 (40.3) | 145 (30.7) | 555 (55.0) | 526 (70.7) |
| Sepsis | 147 (14.7) | 231 (23.1) | 120 (21.9) | 125 (26.5) | 196 (19.4) | 147 (19.8) |
| Aspiration | 96 (9.6) | 149 (14.9) | 84 (15.3) | 72 (15.3) | 58 (5.7) | 49 (6.6) |
| Trauma | 36 (3.6) | 74 (7.4) | 45 (8.2) | 59 (12.5) | 31 (3.1) | 6 (0.8) |
| Other | 71 (7.1) | 73 (7.3) | 79 (14.4) | 71 (15.0) | 170 (16.8) | 16 (2.2) |
| Severity of Illness* | 73.0 (59.0 - 89.0) | 78.0 (62.0 - 94.0) | 78.0 (64.0 - 93.0) | 83.0 (70.0 - 97.0) | 63.0 (50.2 - 75.0) | 76.0 (61.0 - 92.0) |
| Vasopressors - no. (%) | 489 (48.9) | 397 (40.5) | 156 (29.3) | 147 (31.3) | 734 (72.7) | 395 (54.2) |
| Laboratory tests |  |  |  |  |  |  |
| White blood cell count, 10<sup>9</sup>/L | 12.0 (7.8 - 16.7) | 11.8 (7.2 - 17.1) | 11.6 (7.7 - 15.7) | 11.5 (7.5 - 16.2) | --- | 13.9 (8.7 - 20.0) |
| Platelets, 10<sup>9</sup>/L | 169 (108 - 241) | 183 (106 - 258) | 157 (83 - 247) | 135 (80 - 211) | 175 (106 - 263) | 167 (96 - 247) |
| Creatinine, mg/dL | 1.2 (0.8 - 2.0) | 1.0 (0.7 - 1.5) | 1.0 (0.7 - 1.7) | 1.1 (0.8 - 1.7) | 1.3 (0.8 - 2.2) | 1.0 (0.7 - 1.7) |
| Bilirubin, mg/dL | 0.8 (0.5 - 1.4) | 0.8 (0.5 - 1.6) | 0.8 (0.5 - 1.5) | 1.0 (0.6 - 2.1) | 0.8 (0.4 - 1.5) | 0.8 (0.5 - 1.4) |
| Arterial blood gas |  |  |  |  |  |  |
| pH* | 7.36 (7.30 - 7.42) | 7.37 (7.30 - 7.43) | 7.40 (7.34 - 7.44) | 7.41 (7.35 - 7.45) | 7.28 (7.19 - 7.36) | 7.37 (7.31 - 7.42) |
| PaO<sub>2</sub>, mmHg | 83 (68 - 108) | 79 (67 - 100) | 77 (67 - 93) | 76.5 (67 - 93) | 112 (81 - 155) | 83 (69 - 103) |
| PaO<sub>2</sub> / FiO<sub>2</sub> | 125 (86 - 178) | 118 (80 - 163) | 134 (96 - 185) | 112 (75 - 158) | 112 (81 - 155) | 133 (89 - 178) |
| PaCO<sub>2</sub>, mmHg | 38 (34 - 45) | 39 (34 - 45) | 38 (33 - 43) | 36 (31 - 41) | 50 (42 - 62) | 39 (34 - 45) |
| Bicarbonate, mmol/L | 21.0 (18.0 - 25.0) | 21.0 (17.4 - 25.0) | 22.0 (18.0 - 26.0) | 22.0 (18.0 - 25.0) | 22.9 (19.4 - 26.3) | 22.0 (18.0 - 25.0) |
| Ventilatory variables |  |  |  |  |  |  |
| Tidal volume, mL | 410 (360 - 470) | 450 (400 - 510) | 500 (420 - 600) | 700 (600 - 750) | 350 (308 - 400) | 400 (350 - 460) |
| Per PBW, mL/kg PBW | 6.3 (6.0 - 7.3) | 7.1 (6.1 - 8.1) | 7.9 (6.6 - 9.4) | 10.2 (9.0 - 11.3) | 5.9 (5.1 - 6.1) | 6.2 (6.0 - 7.1) |
| Plateau pressure, cmH<sub>2</sub>O | 24.0 (20.0 - 27.0) | 26.0 (22.0 - 30.0) | 26.0 (22.0 - 31.0) | 29.0 (24.8 - 34.0) | 26.0 (22.0 - 29.0) | 24.0 (19.0 - 28.0) |
| PEEP, cmH<sub>2</sub>O | 10 (5 - 12) | 10 (5 - 12) | 10 (5 - 12) | 8 (5 - 10) | 12 (10 - 14) | 10 (5 - 11) |
| FiO<sub>2</sub> | 0.60 (0.50 - 0.80) | 0.60 (0.50 - 0.80) | 0.60 (0.50 - 0.80) | 0.60 (0.50 - 0.74) | 0.70 (0.60 - 1.00) | 0.60 (0.40 - 0.70) |
| Clinical outcomes |  |  |  |  |  |  |
| 28-day mort. - no. (%) | --- | --- | --- | --- | 594 (58.8) | --- |
| 60-day mort. - no. (%) | 227 (22.7) | 268 (26.9) | 144 (26.2) | 141 (30.1) | --- | 199 (26.7) |
| 90-day mort. - no. (%) | 233 (23.3) | 283 (28.6) | 148 (27.5) | 143 (30.8) | --- | 204 (27.4) |
| Ventilator-free days, day 28 | 20.0 (0.0 - 24.0) | 17.0 (0.0 - 23.0) | 18.0 (0.0 - 24.0) | 13.0 (0.0 - 23.0) | 0.0 (0.0 - 13.0) | 20.0 (0.0 - 25.0) |
| Ventilator days in survivors | 7.0 (4.0 - 13.0) | 8.0 (5.0 - 16.0) | 8.0 (4.0 - 14.0) | 8.0 (4.0 - 15.0) | 13.0 (8.0 - 20.0) | 6.0 (4.0 - 11.0) |
| Data are median (quartile 25<sup>th</sup> - quartile 75<sup>th</sup>) or N (%)<br>Abbreviations: 28-day mort. is 28-day mortality, 60-day mort. is 60 days mortality, and 90-day mort. is 90-day mortality.<br>* Except for ART, that uses SAPS-3, all studies use APACHE-IV |  |  |  |  |  |  |

### Predictor variables and model selection

The correlation between the 15 variables selected for clustering is shown in **eTable 3**. The strongest correlation was between PEEP and FiO_2_ (*r* = 0.49). The comparison of the 10 models regarding the optimal number of clusters based on both the Elbow method and the Calinski-Harabasz index is shown in **eFigure 1**. In all models and methods, two clusters were a better fit than a higher number of clusters.

Across the ten models, absolute mortality difference between cluster 1 and cluster 2 ranged from 3.9% to 13.1% for the FACTT study and between 0.1% to 8.1% for EDEN (**Table 2**). The models with the highest 60-day absolute mortality separation between the clusters for each of the two trials in the training set were then further evaluated. Models 6, 5, and 8 were consistently amongst the models with highest separation (**Table 2**). Model 8 was selected for further investigation, as it the fewest variables (**eTable 4**).

**Table 2.** Absolute 60-day Mortality Difference Among Clusters per Trial and Model.

|  |  |  |  |  |  |
| --- | --- | --- | --- | --- | --- |
| **FACTT trial**<br>**(n = 998)** |  |  | **EDEN trial**<br>**(n = 1000)** |  |  |
| **Model** | **Patients scored** | **Mortality difference among clusters** | **Model** | **Patients scored** | **Mortality difference among clusters** |
| 6 | 93.5% | 13.1% | 7 | 77.7% | 8.1% |
| 2 | 57.4% | 12.5% | 8 | 77.7% | 8.1% |
| 5 | 65.5% | 12.2% | 6 | 84.1% | 6.7% |
| 8 | 70.2% | 11.6% | 5 | 71.7% | 6.5% |
| 7 | 70.2% | 11.4% | 9 | 84.7% | 6.1% |
| 1 | 57.4% | 11.2% | 3 | 77.7% | 4.4% |
| 4 | 70.2% | 10.6% | 4 | 77.7% | 4.0% |
| 9 | 93.5% | 10.4% | 2 | 57.7% | 3.9% |
| 3 | 70.2% | 10.1% | 10 | 87.3% | 2.8% |
| 10 | 98.8% | 3.9% | 1 | 57.7% | 0.1% |

### Clinical characteristics of each cluster

Based on model 8, only nine clinical and laboratory variables were needed to identify the two distinct clusters in ARDS patients, namely: heart rate, mean arterial pressure, respiratory rate, bilirubin, bicarbonate, creatinine, PaO_2_, arterial pH, and FiO_2_. For each variable in the model, opposing measurements could be observed for each cluster (**Figure 1** and **eFigure 2**). For the ARDSnet trials, the incidence of cluster 1 patients varied from 57.8% (EDEN) to 73.6% (ARMA), and 41.5% of ART patients were part of cluster 1. Across all trials, patients in cluster 2 had higher severity of illness, rate of vasopressor, heart rate, respiratory rate, creatinine, and bilirubin, as well as lower platelets, pH, BUN, and bicarbonate compared to patients in cluster 1 (**eTable 5, 6** and **7**). In addition, 28-, 60-, and 90-day mortality rate was higher in patients in cluster 2 in all trials (**Table 3**). Likewise, for each trial, ventilator-free days at day 28 was lower in patients in cluster 2 compared to cluster 1, and duration of ventilation in survivors was longer in cluster 1.

**Figure 1.**
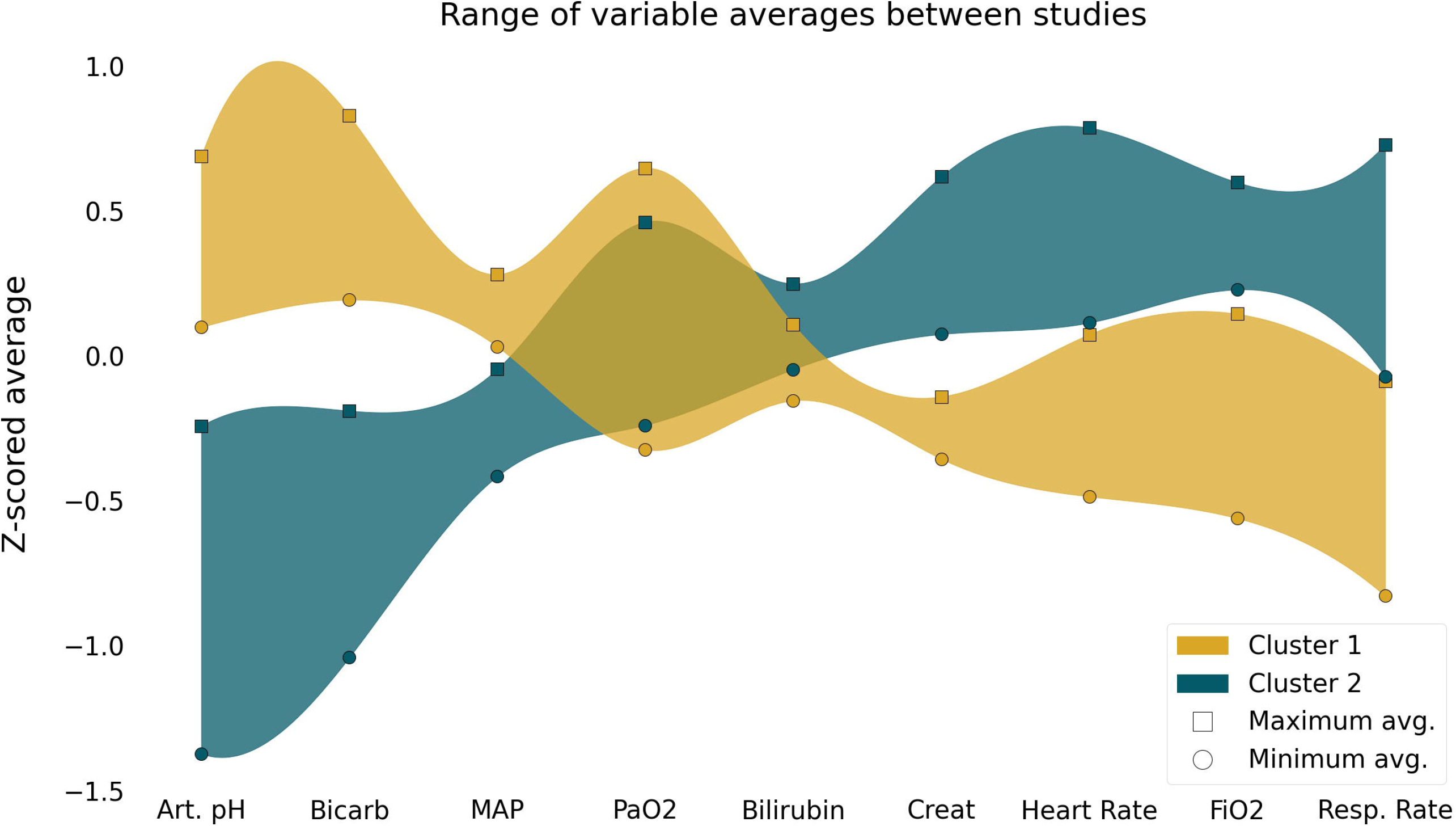
Differences of the Variables Included in the Cluster Algorithm Among Clusters. Square symbols represent the study with the highest mean z score for each phenotype; Circles represent the study with the lowest mean z score for each phenotype. The colored bands are exclusively to help visualize the opposite trends of the variables on the different clusters; Art.pH: arterial pH; Bicarb: bicarbonate; MAP: mean arterial pressure; Creat: creatinine; Resp.Rate: respiratory rate

**Table 3.**
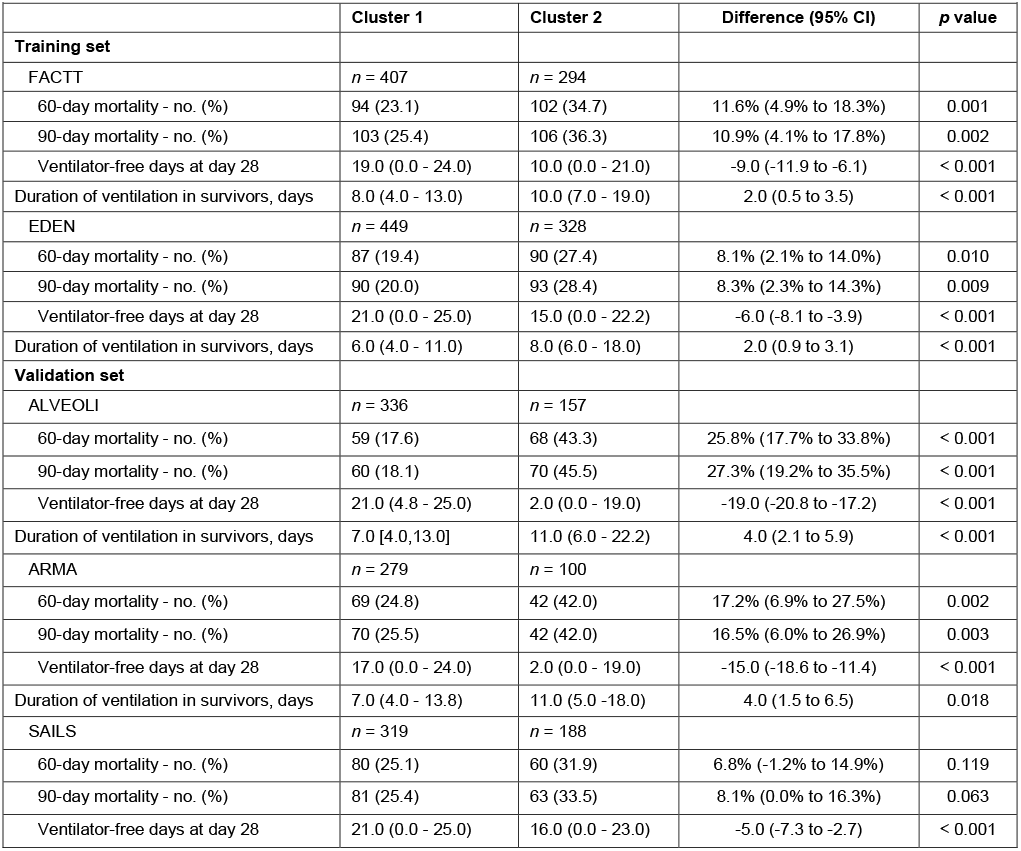
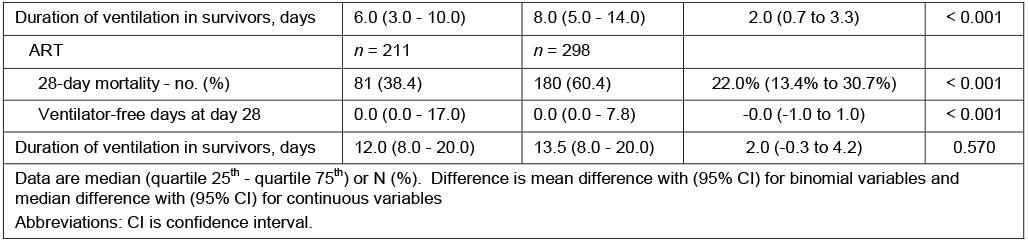
Clinical Outcomes According to Clusters in Each Trial.

### Identification of Subphenotypes

After comparing the clinical characteristics of the clusters, each cluster was assigned to represent a distinct subphenotype of ARDS, with patients in cluster 1 assigned to subphenotype A, and patients in cluster 2 assigned to subphenotype B. Using blood biomarker information available for a subset of patients from both ARMA and ALVEOLI, subphenotype B showed increased levels of pro-inflammatory markers when compared to subphenotype A (**Figure 2** and **eTables 8 and 9**).

**Figure 2.**
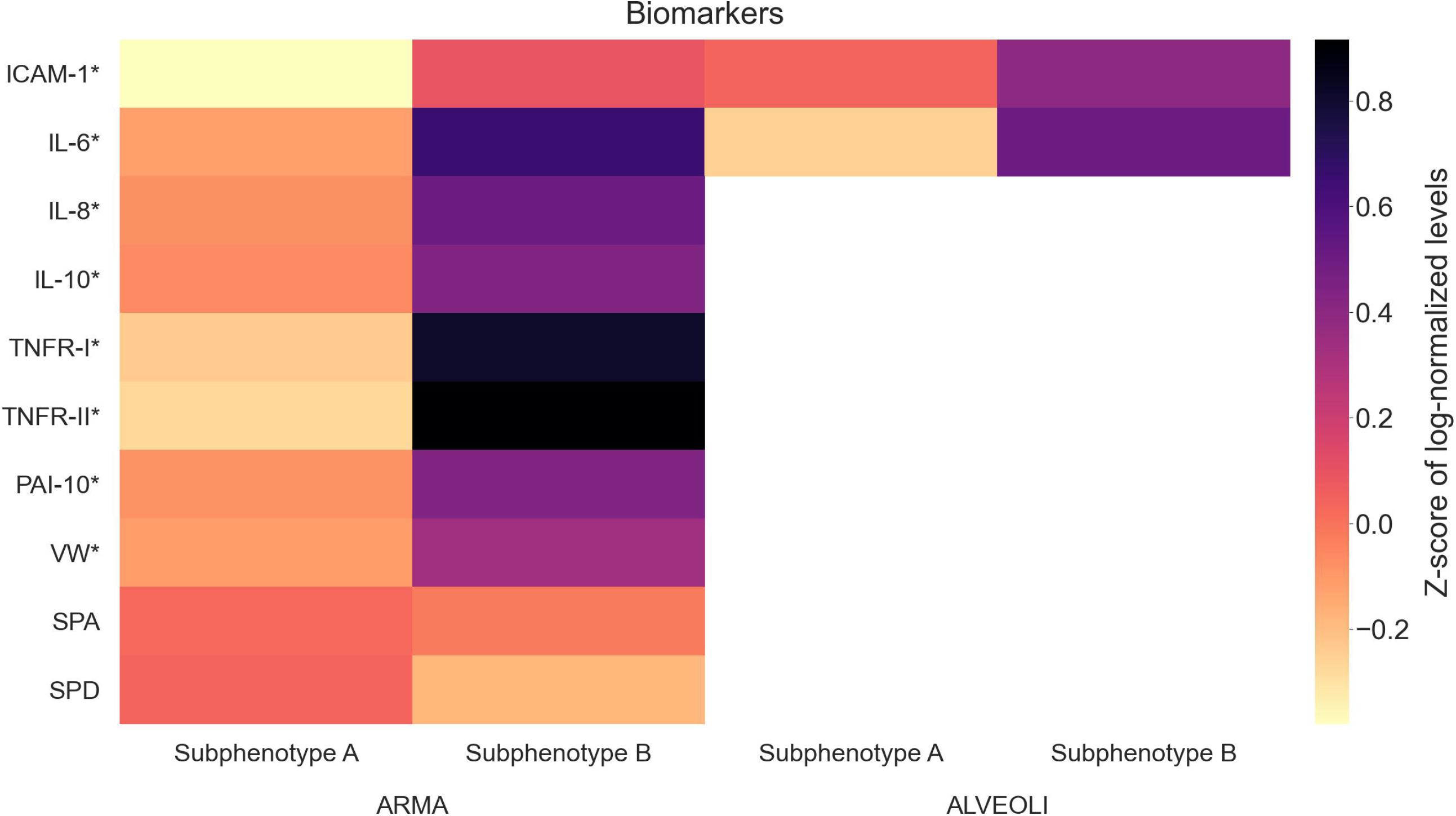
Heat Map of the Biomarkers Available for the ARMA and ALVEOLI Trials. For better visualization and due to difference in scales, the values were log-normalized and z-scored. Subphenotypes A and B are shown separately to highlight their differences.

## DISCUSSION

This study successfully demonstrated that nine easily obtainable clinical variables can identify two distinct ARDS subphenotypes with different clinical and biologic characteristics as well as outcomes across the test and validation cohorts. There was good generalizability amongst diverse populations from multiple validation datasets with temporal and geographical differences.

It is understandable that researchers feel compelled to use as much information as possible to build robust models. This is supportable for two main reasons: (1) the well-known heterogeneity of complex syndromes such as ARDS and (2) the abundance of highly granular clinical data generated by electronic health records (EHRs). It is anticipated that analyzing this vast amount of data will provide new knowledge regarding disease mechanisms by enabling researchers to find plausible hidden patterns within the data [29]. However, this data-heavy approach has the potential drawback of using predictors which are not generally obtained in a time window prior to intervention, or worse yet, using variables that are not part of the routine standard of care for patients. The rationale of using fewer and easy to collect clinical variables is not new in the field of critical care. Prognostic models have already shown that it is indeed feasible to create meaningful models using fewer predictors [30,31].

Our initial choices to define variables commonly found in the EHR at ARDS diagnosis was inspired by a recent report from the World Health Organization (WHO) which showed an enormous discrepancy of medical devices availability in a survey across 135 countries [29]. Recognizing this inconsistency is essential for widespread implementation of machine learning models regardless of varying availability of resources across countries and health systems [29]. The aim is to provide clinically relevant information within a defined and short time period that might impact the delivery of effective interventions to the right patient population and to as many patients as possible [29].

Recently, Sinha *et al*. developed supervised-learning gradient boosted classifier models trained using 24 or 14 readily available clinical data elements to reproduce biomarker-derived subphenotypes which were previously identified by Calfee *et al*. [17]. Unlike Sinha *et al*., who predicted previously identified subphenotypes, our study has identified two subphenotypes *de novo* using a small set of clinical variables.

Although the subphenotypes that we have identified and those that have been previously published look similar, our work is distinct from previous studies in several ways. We employed different training and validation datasets and also utilized a different and well-established unsupervised learning technique. Moreover, we utilized a process for selecting predictors which is not comparable to previous studies. Acknowledging these differences is crucial. It would not be unexpected to assume that these deviations would be relevant enough to produce different subphenotypes [32]. However, the clinical, laboratory characteristics, and the clinical outcomes of our subphenotypes show that they are remarkably similar to subphenotypes found in previous papers, regardless of methodological differences.

At this point it is not possible to go beyond this comparative analysis, as there is no gold standard definition of ARDS subphenotypes [32]. Nonetheless, our work does provide robust evidence that ARDS does indeed have two subphenotypes that can be systematically identified, despite major differences in population assessed and methodological approach used compared with previous studies. It also reinforces that we should continue to explore the underlying biological pathways of such subphenotypes to find responders to new or previously tested therapies.

Our study has several strengths. First, it is the largest cohort of patients that has been studied to develop distinct subphenotypes of ARDS patients. Moreover, our validation cohort included patients from the ART trial, allowing us to validate our model in the contemporaneous population of a large international randomized clinical trial in addition to the ARDSnet studies used in other subphenotyping studies. Second, our subphenotyping model was developed exclusively on the training set and then validated across multiple separate datasets. Nevertheless, similar separation in mortality was seen between the two subphenotypes across all trials. Third, we used the K-means algorithm to identify our subphenotypes, and the results obtained with this technique can be easily interpreted by clinicians and implemented in clinical practice. Lastly, this is the first phenotyping study that has used easily available clinical variables to identify ARDS phenotypes *de novo*, which allows for early identification of these patients in the clinical care at the bedside. Using this algorithm with a small number of routinely collected variables could enable our model to be applied in trials that either retrospectively or prospectively assess interventions targeted to each subphenotype. Future work should analyze previous trials to identify possible differential treatment response for the subphenotypes of ARDS patients identified in this study.

This study also has limitations. First, we have developed our models exclusively on patients enrolled in clinical trials. Due to the strict inclusion and exclusion criteria of these clinical trials, the generalizability of these results needs to be evaluated in unselected ARDS populations. Although there are clear clinical and biomarker differences between the identified subphenotypes, the model’s clinical utility needs to be prospectively validated and further investigated. Additionally, our biomarker analysis is limited to those patients in which the data was made publicly available by the study authors. Lastly, K-means clustering does not handle missing data, and no approach was used to impute missing values. However, the extremely low rate of missingness in our study makes this issue less relevant.

## CONCLUSIONS

This study confirms the existence of two distinct subphenotypes in ARDS patients using a novel clustering model on routinely collected clinical data. This work may allow for easier identification of ARDS subphenotypes to facilitate implementation of precision clinical trial enrollment and development of targeted therapies in a variety of settings without the added burdens of biomarker evaluation.

## Supporting information

TRIPOD

## Data Availability

Data from the ARDSnet studies (EDEN, FACTT, ARMA, ALVEOLI) is publicly available from the NHLBI ARDS Network (NHLBI ARDS Network) and data from the ART trial can be requested from study authors.

## DECLARATIONS

### Funding

This research received no specific grant from any funding agency in the public, commercial or not-for-profit sectors.

### Competing Interest

AD, MS, FGZ, ABC, ISM, DMP, LNL declare no relevant financial conflicts of interest. RK, EVA, LB, JO, DR and ROD are employees of Endpoint Health, Inc. ASN reported receiving personal fees from Dräger unrelated to the submitted work.

### Ethics Approval

All patients provided informed consent in the original trials. This secondary analysis study was determined to be not human subject research and was this exempt from IRB review by WIRB. WIRB Work Order #1-1228617-1

## Author Contributions

AD, RK, EVA, LB participated in study design and analysis, drafted, and revised the manuscript, and are the guarantor of the document. MS, DR, JO, FGZ, ABC, ISM, DMP, LNL, and ASN participated in interpretation of data analysis, drafted the manuscript, and revised it for critically important intellectual content. ROD participated in the study design, analysis, interpretation of data analysis, and final revision of the manuscript content.

### Twitter

@msiuba, @f_g_zampieri, @rod_deliberato, @a_serpaneto, @l_bulgarelli, @endpointhealth

## Notes

### Author Declarations

All patients provided informed consent in the original trials. This secondary analysis study was determined to be not human subject research and was thus exempt from IRB review by WIRB. Specifically de-identified samples and accompanying clinical information that were collected for another purpose will be used for this research. WIRB Work Order #1-1228617-1

