## Supplemental file for "Identification of Acute Respiratory Distress Syndrome subphenotypes denovo using routine clinical data: a retrospective analysis of ARDS clinical trials"

ONLINE SUPPLEMENT

**Additional Methods**

*Number of clusters*

The optimal number of clusters was chosen according to two criteria: (1) Elbow method, by selecting a number of clusters that if further increased will result in only a small increase in performance and possibly cause overfit, hence this number is commonly referenced as to being in the “elbow” of the curve (**eFigure 1**); and (2) Calinski-Harabasz index, consisting of the ratio of *within* to *between* cluster dispersion; higher scores are indication of dense and well separated clusters (**e-Figure 1**).

*Ventilator-free days*

Ventilator free days for ALVEOLI, EDEN, FACTT, and SAILS were calculated according to the methods outlined by Yehya et al (1). Briefly, patients who died at any time in the 28 days were assigned 0 ventilator-free days. For survivors, the number of ventilator-free days was calculated based on the date of the final successful extubation; reintubations before the final extubation were not counted toward ventilator-free days. All days after a patient was discharged home up to the 28^th^ day with unassisted breathing were assumed to be ventilator-free days.

| **eTable 1 - Percentage of missing data in the routinely collected variables, closest randomization, on EDEN and FACTT trials.** | | |
| --- | --- | --- |
|  | **EDEN**  **(*n* = 1000)** | **FACTT**  **(*n* = 999)** |
| Age | 0.0 | 0.0 |
| Gender | 0.0 | 0.0 |
| Arterial pH | 2.8 | 3.9 |
| Bicarbonate | 0.2 | 1.5 |
| Bilirubin | 8.1 | 26.8 |
| Creatinine | 0.0 | 0.0 |
| FiO_2_ | 0.8 | 0.6 |
| Heart Rate | 0.0 | 0.1 |
| Height | 0.1 | 0.9 |
| Mean Arterial Pressure | 12.1 | 0.8 |
| PaCO_2_ | 2.8 | 3.9 |
| PaO_2_ | 0.2 | 4.0 |
| Positive end-expiratory pressure | 1.0 | 0.3 |
| Platelets | 8.1 | 6.0 |
| Plateau pressure | 32.3 | 30.9 |
| Respiratory rate | 0.6 | 0.4 |
| Tidal volume | 15.3 | 12.1 |
| Tidal volume per PBW | 15.4 | 12.8 |

| **eTable 2 - Plausible physiological ranges for clinical measurements, closest to time of randomization** | | |
| --- | --- | --- |
| **Variables** | **Lower Limit** | **Upper Limit** |
| Age (years) | 16 | 89 |
| Arterial pH | 6.65 | 7.80 |
| Bicarbonate (mEq/L) | 1 | 50 |
| Bilirubin (mg/dL) | 0.1 | 50 |
| Creatinine (mg/dL) | 0.1 | 20 |
| FiO2 | 0.21 | 1 |
| Heart Rate (beats per minute) | 20 | 300 |
| Height (cm) | 120 | 220 |
| Mean arterial pressure (mmHg) | 10 | 400 |
| PaCO2 (mmHg) | 20 | 120 |
| PaO2 / FiO2 | 0 | 500 |
| PaO2 (mmHg) | 30 | 500 |
| PEEP (cm H20) | 0 | 60 |
| Platelets (thousands) | 1 | 1000 |
| Plateau Pressure (cm H20) | 10 | 50 |
| Respiratory Rate (resp per minute) | 1 | 100 |
| Tidal Volume (cm H20) | 100 | 1400 |

| **eTable 3 - Correlation among fifteen routinely collected variables, close to the time of randomization.** | | | | | | | | | | | | | | | |
| --- | --- | --- | --- | --- | --- | --- | --- | --- | --- | --- | --- | --- | --- | --- | --- |
|  | **Age** | **pH** | **HCO_3_** | **Bili** | **Creat** | **FiO_2_** | **Gender** | **HR** | **MAP** | **PaCO_2_** | **PaO_2_** | **PEEP** | **Plat** | **RR** | **V_T_/PBW** |
| Age | 1.00 | 0.06 | -0.04 | -0.02 | 0.11 | -0.13 | 0.00 | -0.27 | -0.12 | -0.11 | -0.06 | -0.22 | 0.00 | -0.11 | 0.03 |
| pH | 0.06 | 1.00 | 0.40 | -0.04 | -0.16 | -0.26 | -0.01 | -0.18 | 0.15 | -0.39 | 0.00 | -0.20 | 0.05 | -0.21 | 0.07 |
| HCO_3_ | -0.04 | 0.40 | 1.00 | -0.08 | -0.28 | -0.05 | -0.02 | -0.18 | 0.08 | 0.44 | 0.02 | -0.05 | 0.15 | -0.24 | -0.07 |
| Bili | -0.02 | -0.04 | -0.08 | 1.00 | 0.06 | -0.03 | -0.04 | 0.01 | -0.04 | -0.01 | 0.03 | 0.01 | -0.20 | 0.04 | -0.01 |
| Creat | 0.11 | -0.16 | -0.28 | 0.06 | 1.00 | -0.04 | -0.08 | -0.04 | -0.01 | -0.14 | 0.00 | -0.06 | -0.12 | 0.02 | 0.00 |
| FiO_2_ | -0.13 | -0.26 | -0.05 | -0.03 | -0.04 | 1.00 | 0.03 | 0.13 | -0.06 | 0.18 | 0.11 | 0.49 | 0.06 | 0.21 | -0.02 |
| Gender | 0.00 | -0.01 | -0.02 | -0.04 | -0.08 | 0.03 | 1.00 | -0.03 | -0.05 | -0.04 | -0.06 | 0.02 | 0.09 | 0.09 | 0.19 |
| HR | -0.27 | -0.18 | -0.18 | 0.01 | -0.04 | 0.13 | -0.03 | 1.00 | -0.02 | 0.03 | -0.04 | 0.12 | -0.05 | 0.22 | 0.08 |
| MAP | -0.12 | 0.15 | 0.08 | -0.04 | -0.01 | -0.06 | -0.05 | -0.02 | 1.00 | -0.03 | 0.01 | -0.01 | 0.06 | -0.04 | 0.00 |
| PaCO_2_ | -0.11 | -0.39 | 0.44 | -0.01 | -0.14 | 0.18 | -0.04 | 0.03 | -0.03 | 1.00 | -0.04 | 0.17 | 0.11 | -0.05 | -0.17 |
| PaO_2_ | -0.06 | 0.00 | 0.02 | 0.03 | 0.00 | 0.11 | -0.06 | -0.04 | 0.01 | -0.04 | 1.00 | -0.09 | -0.04 | -0.09 | 0.03 |
| PEEP | -0.22 | -0.20 | -0.05 | 0.01 | -0.06 | 0.49 | 0.02 | 0.12 | -0.01 | 0.17 | -0.09 | 1.00 | 0.00 | 0.33 | -0.15 |
| Plat | 0.00 | 0.05 | 0.15 | -0.20 | -0.12 | 0.06 | 0.09 | -0.05 | 0.06 | 0.11 | -0.04 | 0.00 | 1.00 | -0.05 | 0.03 |
| RR | -0.11 | -0.21 | -0.24 | 0.04 | 0.02 | 0.21 | 0.09 | 0.22 | -0.04 | -0.05 | -0.09 | 0.33 | -0.05 | 1.00 | -0.31 |
| V_T_/PBW | 0.03 | 0.07 | -0.07 | -0.01 | 0.00 | -0.02 | 0.19 | 0.08 | 0.00 | -0.17 | 0.03 | -0.15 | 0.03 | -0.31 | 1.00 |
| Data are Pearson correlation coefficients.  Abbreviations: Bili denotes bilirubin, Creat is creatinine, HR is heart rate, MAP is mean arterial pressure, PEEP is positive end-expiratory pressure, Plat is platelets, RR is respiratory rate and V_T_/PBW is tidal volume per predicted body weight. | | | | | | | | | | | | | | | |

| **eTable 4 - List of variables in each model assessed** | | | | | | | | | | | | | | | | |
| --- | --- | --- | --- | --- | --- | --- | --- | --- | --- | --- | --- | --- | --- | --- | --- | --- |
| **Model** | **Demographics** | | **Arterial Blood Gases** | | | **Laboratory Values** | | | | | **Vital Signs** | | | **Ventilator Variables** | | |
|  | **Age** | **Gender** | **pH** | **PaO_2_** | **PaCO_2_** | | **Creat** | **Bili** | **HCO_3_** | **Plat** | **MAP** | **RR** | **HR** | **FiO2** | **PEEP** | **V_T_/PBW** |
| 1 | X | X | X | X | X | X | | X | X | X | X | X | X | X | X | X |
| 2 |  |  | X | X | X | X | | X | X | X | X | X | X | X | X | X |
| 3 | X | X | X | X | X | X | | X | X |  | X | X | X | X |  |  |
| 4 | X | X | X | X |  | X | | X | X |  | X | X | X | X |  |  |
| 5 |  |  | X | X | X | X | | X | X | X | X | X | X | X |  |  |
| 6 | X | X | X | X |  | X | |  | X |  | X | X | X | X |  |  |
| 7 |  |  | X | X | X | X | | X | X |  | X | X | X | X |  |  |
| 8 |  |  | X | X |  | X | | X | X |  | X | X | X | X |  |  |
| 9 |  |  | X | X | X |  | |  | X |  | X | X | X |  |  |  |
| 10 | X | X |  |  |  |  | |  |  |  | X | X | X |  |  |  |
| Abbreviations: Bili denotes bilirubin, Creat is creatinine, HR is heart rate, MAP is mean arterial pressure, PEEP is positive end-expiratory pressure, Plat is platelets, RR is respiratory rate and V_T_/PBW is tidal volume per predicted body weight. | | | | | | | | | | | | | | | | |

| **eTable 5 - Baseline Characteristics and Clinical Outcomes According to the Clusters and Trials in the Training Set** | | | | | | |
| --- | --- | --- | --- | --- | --- | --- |
|  | **FACTT** | | | **EDEN** | | |
|  | **Cluster 1**  **(*n* = 407)** | **Cluster 2**  **(*n* = 294)** | ***p v*alue** | **Cluster 1**  **(*n* = 449)** | **Cluster 2**  **(*n* = 328)** | ***p v*alue** |
| Age, year* | 50.0 (40.0 - 63.0) | 47.0 (36.0 - 58.0) | 0.002 | 53.0 (44.0 - 63.0) | 51.0 (41.0 - 62.2) | 0.183 |
| Male gender - no. (%) | 223 (54.8) | 151 (51.4) | 0.411 | 233 (51.9) | 168 (51.2) | 0.910 |
| Body mass index, kg/m^2^ | 27.5 (23.3 - 32.1) | 27.4 (23.0 - 32.7) | 0.938 | 29.1 (24.6 - 34.5) | 28.5 (23.4 - 35.1) | 0.476 |
| Caucasian - no. (%) | 269 (66.1) | 177 (60.2) | 0.129 | 349 (81.5) | 237 (75.7) | 0.067 |
| Etiology - no. (%) |  |  | < 0.001 |  |  | 0.003 |
| Pneumonia | 201 (49.4) | 139 (47.3) |  | 296 (65.9) | 217 (66.2) |  |
| Sepsis | 78 (19.2) | 101 (34.4) |  | 50 (11.1) | 60 (18.3) |  |
| Aspiration | 67 (16.5) | 30 (10.2) |  | 45 (10.0) | 27 (8.2) |  |
| Trauma | 24 (5.9) | 8 (2.7) |  | 24 (5.3) | 5 (1.5) |  |
| Other | 37 (9.1) | 16 (5.4) |  | 34 (7.6) | 19 (5.8) |  |
| Prognostic scores |  |  |  |  |  |  |
| APACHE III | 69.0 (56.0 - 84.0) | 91 (76.0 - 105.0) | < 0.001 | 66.0 (54.0 - 79.0) | 84.0 (71.0 - 100.2) | < 0.001 |
| Use of vasopressor - no. (%) | 118 (29.5) | 189 (64.9) | < 0.001 | 187 (41.6) | 209 (63.7) | < 0.001 |
| Vital signs |  |  |  |  |  |  |
| Temperature, ºC | 37.5 (36.8 - 38.2) | 37.6 (37.0 - 38.4) | 0.371 | 37.3 (36.8 - 37.8) | 37.3 (36.7 - 38.1) | 0.212 |
| Heart rate, bpm | 95.0 (81.0 - 110.0) | 114 (102 - 126) | < 0.001 | 89 (77 - 102) | 101 (89 - 116) | < 0.001 |
| Mean arterial Pressure, mmHg | 76.0 (68.0 - 88.0) | 71.0 (65.0 - 80.8) | < 0.001 | 77.0 (68.0 - 84.0) | 71.0 (66.0 - 80.0) | < 0.001 |
| SpO_2_, % | 96 (93 - 98) | 95 (92 - 97) | < 0.001 | 96 (94 - 98) | 95 (92 - 98) | 0.032 |
| Urine output in 24 hours, mL | 1785 (1192 - 2853) | 1370 (842 - 2446) | < 0.001 | 1505 (977 - 2250) | 1165 (566 - 1816) | < 0.001 |
| Laboratory tests |  |  |  |  |  |  |
| Hematocrit, % | 30.0 (26.0 - 33.0) | 30.0 (24.2 - 35.0) | 0.272 | 30.0 (26.0 - 34.0) | 30.0 (26.0 - 35.0) | 0.919 |
| White blood cell count, 10^9^/L | 11.6 (7.3 - 16.3) | 11.7 (5.6 - 17.9) | 0.972 | 11.4 (7.7 - 15.5) | 12.7 (7.7 - 19.0) | 0.019 |
| Platelets, 10^9^/L | 195 (118.5 - 268) | 158 (87 - 237) | < 0.001 | 163 (108 - 241) | 164 (103 - 227) | 0.552 |
| Creatinine, mg/dL | 0.9 (0.7 - 1.3) | 1.4 (1.0 - 2.0) | < 0.001 | 1.0 (0.7 - 1.5) | 1.6 (1.0 - 2.8) | < 0.001 |
| Bilirubin, mg/dL | 0.7 (0.5 - 1.3) | 0.9 (0.5 - 2.0) | 0.003 | 0.8 (0.5 - 1.3) | 0.8 (0.5 - 1.7) | 0.128 |
| Arterial blood gas |  |  |  |  |  |  |
| pH* | 7.41 (7.36 - 7.45) | 7.29 (7.23 - 7.35) | < 0.001 | 7.40 (7.35 - 7.44) | 7.30 (7.24 - 7.35) | < 0.001 |
| PaO_2_, mmHg | 78 (68 - 100) | 78 (65 - 99) | 0.240 | 83 (70 - 107) | 81 (67 - 107) | 0.416 |
| PaO_2_ / FiO_2_ | 132 (92 - 173) | 89 (65 - 126) | < 0.001 | 133 (98 - 193) | 101 (73 - 162) | < 0.001 |
| PaCO_2_, mmHg | 39 (34 - 44) | 38.5 (33 - 47.9) | 0.877 | 38 (34 - 44) | 38 (33 - 46) | 0.55 |
| Bicarbonate, mmol/L | 24.0 (21.0 - 27.0) | 17.0 (14.0 - 20.0) | < 0.001 | 23.0 (21.0 - 26.0) | 18.5 (15.0 - 21.0) | < 0.001 |
| Ventilatory variables |  |  |  |  |  |  |
| Tidal volume, mL | 450 (400 - 530) | 450 (382 - 500) | 0.009 | 420 (356 - 487) | 400 (350 - 450) | 0.032 |
| Per PBW, mL/kg PBW | 7.1 (6.3 - 8.4) | 7.0 (6.0, 8.0) | 0.058 | 6.3 (6.0 - 7.5) | 6.1 (6.0 - 7.3) | 0.079 |
| Plateau pressure, cmH_2_O | 25.0 (20.0 - 29.0) | 28.0 (24.0 - 32.0) | < 0.001 | 23.0 (19.0 - 27.0) | 24.0 (21.0 - 28.0) | 0.004 |
| PEEP, cmH_2_O | 8 (5 - 10) | 10 (8 - 14) | < 0.001 | 10 (5 - 10) | 10 (8 - 14) | < 0.001 |
| Respiratory rate, breaths/min | 22 (18 - 27) | 30 (24 - 35) | < 0.001 | 22 (19 - 26) | 30 (25 - 35) | < 0.001 |
| FiO_2_ | 0.50 (0.40 - 0.70) | 0.80 (0.60 - 1.00) | < 0.001 | 0.60 (0.45 - 0.70) | 0.80 (0.60 - 1.00) | < 0.001 |
| Data are mean ± standard deviation, median (quartile 25^th^ - quartile 75^th^) or N (%)  Abbreviations: APACHE denotes Acute Physiology and Chronic Health Evaluation, V_T_/PBW denotes tidal volume per predicted body weight. | | | | | | |

| **eTable 6 - Baseline Characteristics and Clinical Outcomes According to the Clusters and Two Trials in the Validation Set** | | | | | | |
| --- | --- | --- | --- | --- | --- | --- |
|  | **ALVEOLI** | | | **ARMA** | | |
|  | **Cluster 1**  **(*n* = 336)** | **Cluster 2**  **(*n* = 157)** | ***p v*alue** | **Cluster 1**  **(*n* = 279)** | **Cluster 2**  **(*n* = 100)** | ***p v*alue** |
| Age, year* | 53.0 (39.0 - 66.2) | 46.0 (37.0 - 60.0) | 0.007 | 49.0 (37.0 - 64.0) | 47.5 (36.0 - 61.0) | 0.180 |
| Male gender - no. (%) | 188 (56.0) | 86 (54.8) | 0.883 | 169 (60.6) | 61 (61.0) | 0.965 |
| Body mass index, kg/m^2^ | 27.0 (22.9 - 31.1) | 25.2 (21.7 - 30.2) | 0.050 | 25.8 (23.0 - 30.2) | 24.4 (21.5 - 29.7) | 0.057 |
| Caucasian - no. (%) | 263 (78.3) | 102 (65.0) | 0.002 | 220 (78.9) | 65 (65.0) | 0.009 |
| Etiology - no. (%) |  |  | 0.001 |  |  | < 0.001 |
| Pneumonia | 130 (38.7) | 66 (42.0) |  | 83 (29.7) | 30 (30.0) |  |
| Sepsis | 63 (18.8) | 50 (31.8) |  | 64 (22.9) | 43 (43.0) |  |
| Aspiration | 55 (16.4) | 19 (12.1) |  | 44 (15.8) | 14 (14.0) |  |
| Trauma | 33 (9.8) | 5 (3.2) |  | 43 (15.4) | 4 (4.0) |  |
| Other | 55 (16.4) | 17 (10.8) |  | 45 (16.1) | 9 (9.0) |  |
| Prognostic scores |  |  |  |  |  |  |
| APACHE III | 71. (59.0 - 83.0) | 93.0 (80.0 - 110.0) | < 0.001 | 77.0 (66.0 - 90.5) | 97.0 (81.8 (110.0) | < 0.001 |
| Use of vasopressor - no. (%) | 65 (20.1) | 80 (51.3) | < 0.001 | 77 (27.6) | 52 (52.5) | < 0.001 |
| Vital signs |  |  |  |  |  |  |
| Temperature, ºC | 37.6 (37.1 - 38.2) | 37.7 (36.9 - 38.3) | 0.778 | 37.6 (37.1 - 38.1) | 37.6 (36.8 - 38.4) | 0.803 |
| Heart rate, bpm | 97.5 (83.0 - 109) | 111.0 (97.0 - 126) | < 0.001 | 101.0 (89.0 - 112.5) | 118 (105.0 - 128.0) | < 0.001 |
| Mean arterial Pressure, mmHg | 77.3 (77.0 - 87.3) | 73.3 (65.0 - 80.3) | < 0.001 | 78.0 (70.7 - 88.0) | 70.5 (64.9 - 80.4) | < 0.001 |
| SpO_2_, % | 96 (94 - 97) | 95 (92 - 97) | 0.005 | 95 (93 - 98) | 95.5 (93 - 97) | 0.799 |
| Urine output in 24 hours, mL | 2065 (1355 - 3255) | 1433 (569 - 2189) | < 0.001 | 2100 (1375 - 3096) | 1525 (816 - 2650) | 0.001 |
| Laboratory tests |  |  |  |  |  |  |
| Hematocrit, % | 31.0 (28.0 - 34.0) | 31.0 (27.0 - 35.0) | 0.617 | 30.0 (28.0 - 33.0) | 31.0 (28.0 - 34.0) | 0.299 |
| White blood cell count, 10^9^/L | 11.7 (8.1 - 15.3) | 10.7 (6.4 - 15.8) | 0.166 | 11.9 (7.7 - 16.7) | 9.8 (5.4 - 16.7) | 0.057 |
| Platelets, 10^9^/L | 173 (94 - 266) | 141 (57 - 214) | 0.001 | 139 (80 - 212) | 125 (72 - 196) | 0.260 |
| Creatinine, mg/dL | 0.9 (0.7 - 1.3) | 1.5 (0.9 - 3.0) | < 0.001 | 1.0 (0.7 - 1.4) | 1.8 (1.2 - 3.2) | < 0.001 |
| Bilirubin, mg/dL | 0.8 (0.5 - 1.4) | 0.9 (0.4 - 1.8) | 0.289 | 1.0 (0.6 - 2.1) | 1.1 (0.7 - 2.7) | 0.106 |
| Arterial blood gas |  |  |  |  |  |  |
| pH* | 7.42 (7.38 - 7.45) | 7.31 (7.24 - 7.36) | < 0.001 | 7.42 (7.38 - 7.47) | 7.33 (7.28 - 7.37) | < 0.001 |
| PaO_2_, mmHg | 78 (68 - 93) | 74 (65 - 92) | 0.082 | 75 (66 - 91) | 81 (68 - 96) | 0.106 |
| PaO_2_ / FiO_2_ | 149 (109 - 192) | 103 (74 - 136) | < 0.001 | 118 (83 - 160) | 99 (68 - 137) | 0.006 |
| PaCO_2_, mmHg | 38 (34 - 43) | 36 (31 - 42) | 0.046 | 37 (31 - 41) | 34 (28.8 - 39.2) | 0.003 |
| Bicarbonate, mmol/L | 24 (21 - 27) | 17 (13 - 20) | < 0.001 | 23 (20 - 26) | 16 (13 - 19) | < 0.001 |
| Ventilatory variables |  |  |  |  |  |  |
| Tidal volume, mL | 500 (437 - 600) | 480 (400 - 572) | 0.002 | 700 (600 - 750) | 700 (550 - 700) | 0.198 |
| Per PBW, mL/kg PBW | 8.0 (6.9 - 9.5) | 7.4 (6.2 - 9.2) | 0.006 | 10.1 (9.2 - 11.1) | 10.6 (9.0 - 11.4) | 0.383 |
| Plateau pressure, cmH_2_O | 25.0 (21.0 - 30.0) | 29.0 (24.0 - 33.0) | < 0.001 | 29.0 (24.0 - 34.0) | 31.0 (27.0 - 36.0) | 0.018 |
| PEEP, cmH_2_O | 10 (5 - 10) | 10 (8 - 14) | < 0.001 | 8 (5 - 10) | 10 (5 - 12) | 0.150 |
| Respiratory rate, breaths/min | 20 (15 - 25) | 30 (24 - 35) | < 0.001 | 18 (14 - 21) | 24 (18.8 - 28) | < 0.001 |
| FiO_2_ | 0.50 (0.44 - 0.65) | 0.75 (0.60 - 1.00) | < 0.001 | 0.60 (0.50 - 0.70) | 0.70 (0.59 - 0.96) | < 0.001 |
| Data are mean ± standard deviation, median (quartile 25^th^ - quartile 75^th^) or N (%)  Abbreviations: APACHE denotes Acute Physiology and Chronic Health Evaluation, V_T_/PBW denotes tidal volume per predicted body weight. | | | | | | |

| **eTable 7 - Baseline Characteristics and Clinical Outcomes According to the Clusters and Two Trials in the Validation Set** | | | | | | |
| --- | --- | --- | --- | --- | --- | --- |
|  | **SAILS** | | | **ART** | | |
|  | **Cluster 1**  **(*n* = 319)** | **Cluster 2**  **(*n* = 188)** | ***p v*alue** | **Cluster 1**  **(*n* = 211)** | **Cluster 2**  **(*n* = 298)** | ***p v*alue** |
| Age, year* | 57.0 (46.0 - 67.0) | 53.5 (39.0 - 65.0) | 0.035 | 54.0 (37.0 - 65.0) | 50.0 (35.2 - 61.0) | 0.075 |
| Male gender - no. (%) | 150 (47.0) | 100 (53.2) | 0.211 | 136 (64.5) | 181 (60.7) | 0.448 |
| Body mass index, kg/m^2^ | 28.5 (23.9 - 34.6) | 29.8 (23.2 - 35.1) | 0.903 | 28.8 (24.6 - 35.6) | 28.4 (25.0 - 31.7) | 0.367 |
| Caucasian - no. (%) | 250 (78.4) | 140 (74.5) | 0.369 | --- | --- | --- |
| Etiology - no. (%) |  |  | 0.709 |  |  | 0.052 |
| Pneumonia | 228 (71.5) | 127 (67.6) |  | 113 (53.6) | 171 (57.4) |  |
| Sepsis | 63 (19.7) | 39 (20.7) |  | 38 (18.0) | 59 (19.8) |  |
| Aspiration | 19 (6.0) | 15 (8.0) |  | 13 (6.2) | 16 (5.4) |  |
| Trauma | 3 (0.9) | 1 (0.5) |  | 10 (4.7) | 2 (0.7) |  |
| Other | 6 (1.9) | 6 (3.2) |  | 37 (17.5) | 50 (16.8) |  |
| Prognostic scores |  |  |  | --- | --- | --- |
| APACHE III | 70.0 (56.0 - 84.0) | 92.0 (75.0 - 105.8) | < 0.001 |  |  |  |
| SAPS III | --- | --- | --- | 62 (50 - 71) | 66 (53 - 75) | 0.010 |
| Use of vasopressor - no. (%) | 150 (47.8) | 142 (78.5) | < 0.001 | 130 (61.6) | 242 (81.2) | < 0.001 |
| Vital signs |  |  |  |  |  |  |
| Temperature, ºC | 37.2 (36.7 - 37.8) | 37.3 (36.7 - 38.0) | 0.346 | --- | --- | --- |
| Heart rate, bpm | 91.0 (80.5 - 103.0) | 102.0 (88.8 - 117.0) | < 0.001 | 90.0 (73.0 - 103.0) | 112.0 (97.2 - 126.0) | < 0.001 |
| Mean arterial Pressure, mmHg | 78.0 (69.5 - 88.0) | 70.0 (63.0 - 78.) | < 0.001 | 80.0 (73.5 - 89.0) | 75.0 (70.0 - 83.0) | < 0.001 |
| SpO_2_, % | 96 (95 - 99) | 96 (93 - 99) | 0.270 | --- | --- | --- |
| Urine output in 24 hours, mL | 1570 (852 - 2383) | 920 (350 - 1665) | < 0.001 | --- | --- | --- |
| Laboratory tests |  |  |  |  |  |  |
| Hematocrit, % | 31 (27 - 35) | 31 (28 - 37) | 0.142 | --- | --- | --- |
| White blood cell count, 10^9^/L | 13.6 (8.5 - 18.1) | 15.4 (9.8 - 23.3) | 0.009 | --- | --- | --- |
| Platelets, 10^9^/L | 164 (96 - 238) | 131 (80 - 223) | 0.032 | 177 (120 - 292) | 169 (90 - 256) | 0.048 |
| Creatinine, mg/dL | 1.0 (0.7 - 1.5) | 1.4 (0.9 - 2.6) | < 0.001 | 1.0 (0.7 - 1.5) | 1.7 (1.0 - 2.8) | < 0.001 |
| Bilirubin, mg/dL | 0.8 (0.5 - 1.4) | 0.8 (0.5 - 1.6) | 0.630 | 0.6 (0.4 - 1.2) | 0.9 (0.4 - 1.7) | 0.002 |
| Arterial blood gas |  |  |  |  |  |  |
| pH* | 7.39 (7.35 - 7.44) | 7.31 (7.24 - 7.35) | < 0.001 | 7.4 (7.3 - 7.4) | 7.2 (7.2 - 7.3) | < 0.001 |
| PaO_2_, mmHg | 82 (68 - 101) | 86 (72 - 111.2) | 0.112 | 118 (82 - 158) | 104 (78 - 152) | 0.065 |
| PaO_2_ / FiO_2_ | 139 (98 - 195) | 107 (74 - 159) | < 0.001 | 118 (82 - 158) | 104 (78 - 152) | 0.065 |
| PaCO_2_, mmHg | 38 (34 - 45) | 38 (32 - 44) | 0.423 | 46 (41 - 56) | 53 (42 - 65) | < 0.001 |
| Bicarbonate, mmol/L | 23 (20 - 26) | 17 (14 - 21) | < 0.001 | 25.2 (22.5 - 28.8) | 20.6 (17.8 - 23.4) | < 0.001 |
| Ventilatory variables |  |  |  |  |  |  |
| Tidal volume, mL | 420 (360 - 480) | 400 (340 - 450) | 0.016 | 360 (320 - 400) | 350 (300 - 397.8) | 0.008 |
| Per PBW, mL/kg PBW | 6.4 (6.0 - 7.3) | 6.1 (5.9 - 7.0) | 0.030 | 6.0 (5.3 - 6.1) | 5.9 (5.1 - 6.1) | 0.034 |
| Plateau pressure, cmH_2_O | 22.0 (18.0 - 27.0) | 25.0 (20.0 - 29.0) | 0.003 | 24.0 (21.0 - 28.0) | 27.0 (23.0 - 30.0) | < 0.001 |
| PEEP, cmH_2_O | 8 (5 - 10) | 10 (8 - 13) | 0.001 | 10 (10 - 14) | 12 (10 - 14) | < 0.001 |
| Respiratory rate, breaths/min | 23 (19 - 27) | 30 (24 - 35) | < 0.001 | 24 (20 - 28) | 30 (24 - 34) | < 0.001 |
| FiO_2_ | 0.50 (0.40 - 0.60) | 0.70 (0.50 - 0.90) | < 0.001 | 0.70 (0.60 - 0.80) | 0.80 (0.70 - 1.00) | < 0.001 |
| Data are mean ± standard deviation, median (quartile 25^th^ - quartile 75^th^) or N (%)  Abbreviations: APACHE denotes Acute Physiology and Chronic Health Evaluation, V_T_/PBW denotes tidal volume per predicted body weight... | | | | | | |

| **eTable 8 - Biomarker levels by study and cluster** | | | | | | | | |
| --- | --- | --- | --- | --- | --- | --- | --- | --- |
|  | **ARMA** | | | | **ALVEOLI** | | | |
|  | **Subphenotype A (*n* = 279)** | **Subphenotype B (*n* = 100)** | **Median Difference**  **(95% CI)** | ***p* value** | **Subphenotype A (*n* = 336)** | **Subphenotype B (*n* = 157)** | **Median Difference (95% CI)** | ***p* value** |
| ICAM-1 | 654.0 (399.0 - 959.4) | 888.0 (550.0 - 1365.3) | 234 (60.3 to 407.8) | 0.002 | 847.9 (585.7 - 1227.1) | 1070.4 (748.2 - 1588.8) | 219.4 (90.4 to 348.4) | < 0.001 |
| IL-6 | 214.0 (91.8 - 553.5) | 966.0 (291.0 - 2200.0) | 749.1 (589.9 to 908.2) | < 0.001 | 182.5 (85.5 - 435.2) | 775.0 (148.0 - 2846.5) | 592 (515.5 to 668.6) | < 0.001 |
| PAI-1 | 65.3 (37.8 - 109.5) | 101.7 (50.8 - 291.6) | 41 (18.3 to 63.7) | 0.001 | Not assessed | Not assessed | --- | --- |
| IL-8 | 46.0 (2.0 - 91.0) | 106.9 (43.8 - 281.4) | 60.9 (35.6 to 86.2) | < 0.001 | Not assessed | Not assessed | --- | --- |
| IL-10 | 16.0 (0.0 - 40.3) | 47.9 (0.0 - 120.7) | 31.9 (20.2 to 43.6) | < 0.001 | Not assessed | Not assessed | --- | --- |
| TNFR-I | 2604.0 (1950.0 - 3777.0) | 6897.0 (3622.5 - 12281.5) | 4293 (3323.6 to 5262.4) | < 0.001 | Not assessed | Not assessed | --- | --- |
| TNFR-II | 6581.0 (4958.0 - 9658.0) | 18611.0 (12262.5 - 35652.0) | 12030 (9577.5 to 14482.5) | < 0.001 | Not assessed | Not assessed | --- | --- |
| SPA | 29.0 (11.8 - 68.0) | 25.0 (10.5 - 40.0) | -4 (-19.9 to 11.9) | 0.398 | Not assessed | Not assessed | --- | --- |
| SPD | 76.0 (36.2 - 145.2) | 59.0 (30.0 - 125.0) | -18 (-52.6 to 16.6) | 0.254 | Not assessed | Not assessed | --- | --- |
| VW | 308.0 (165.5 - 431.0) | 384.0 (246.0 - 549.0) | 76 (-26.5 to 178.5) | 0.045 | Not assessed | Not assessed | --- | --- |
| Data are median (quartile 25^th^ - quartile 75^th^).  Abbreviations: 95%CI denotes 95% confidence interval, ICAM-1 is intercellular adhesion molecule-1, IL-6 is interleukin-6, PAI-1 is plasminogen activator inhibitor-1, IL-8 is interleukin-8, IL-10 is interleukin-10, TNFR-I is tumor necrosis factor receptor 1, TNFR-II is tumor necrosis factor II, SPA is surfact protein A, SPD is surfact Protein D and VW is Von Willebrand factor. | | | | | | | | |

| **eTable 9 - Percentage of missingness in biomarker levels measured on day of randomization, on ARMA and ALVEOLI trials for patients with an assigned subphenotype** | | | | |
| --- | --- | --- | --- | --- |
| **Biomarker** | **ARMA**  **(*n* = 379)** | | **ALVEOLI**  **(*n* = 493)** | |
|  | **Subphenotype A** | **Subphenotype B** | **Subphenotype A** | **Subphenotype B** |
| ICAM-1 | 43% | 31% | 4% | 3% |
| IL-6 | 41% | 33% | 4% | 4% |
| PAI-1 | 42% | 32% | Not assessed | Not assessed |
| IL-8 | 41% | 33% | Not assessed | Not assessed |
| IL-10 | 42% | 33% | Not assessed | Not assessed |
| TNFR-I | 68% | 61% | Not assessed | Not assessed |
| TNFR-II | 68% | 61% | Not assessed | Not assessed |
| SPA | 67% | 61% | Not assessed | Not assessed |
| SPD | 67% | 61% | Not assessed | Not assessed |
| VW | 67% | 61% | Not assessed | Not assessed |
| Abbreviations: ICAM-1 is intercellular adhesion molecule-1, IL-6 is interleukin-6, PAI-1 is plasminogen activator inhibitor-1, IL-8 is interleukin-8, IL-10 is interleukin-10, TNFR-I is tumor necrosis factor receptor 1, TNFR-II is tumor necrosis factor II, SPA is surfact protein A, SPD is surfact Protein D and VW is Von Willebrand factor. | | | | |

**eFigure 1 - Calinski-Harabasz Index and Elbow Method for Each of the 10 Models**

Calinski-Harabasz Index for Training Data

Calinski-Harabasz Index

| Number of Clusters | model_1 | model_2 | model_3 | model_4 | model_5 | model_6 | model_7 | model_8 | model_9 | model_10 |
| --- | --- | --- | --- | --- | --- | --- | --- | --- | --- | --- |
| 2 | 140.994 | 159.52 | 175.508 | 193.148 | 174.903 | 256.553 | 212.252 | 239.431 | 287.835 | 470.596 |
| 3 | 108.544 | 123.558 | 145.884 | 158.272 | 146.904 | 212.965 | 176.855 | 188.563 | 243.696 | 416.168 |
| 4 | 93.2987 | 107.004 | 132.552 | 143.859 | 134.373 | 193.888 | 162.593 | 178.404 | 223.876 | 390.621 |
| 5 | 84.9043 | 98.4188 | 126.848 | 140.808 | 127.416 | 188.497 | 160.198 | 179.961 | 213.943 | 374.456 |

Elbow Criterion for Training Data

Sum of Squared Error

| Number of Clusters | model_1 | model_2 | model_3 | model_4 | model_5 | model_6 | model_7 | model_8 | model_9 | model_10 |
| --- | --- | --- | --- | --- | --- | --- | --- | --- | --- | --- |
| 2 | 16387.4 | 14135.8 | 15851.2 | 14376.7 | 13372.6 | 15496.4 | 12921.8 | 11445.4 | 13735 | 7415.73 |
| 3 | 15471.7 | 13246.2 | 14807 | 13385.4 | 12414.7 | 14300.7 | 11921.3 | 10593.5 | 12520.3 | 6417.2 |
| 4 | 14788.2 | 12577 | 13967.8 | 12575.9 | 11646.5 | 13352.2 | 11105.1 | 9758.62 | 11574.2 | 5696.42 |
| 5 | 14190.9 | 11980.8 | 13191.9 | 11761 | 10983.1 | 12438.5 | 10299.5 | 8935.36 | 10760.5 | 5141.36 |

**eFigure 2 - Variable Averages for Each Study**

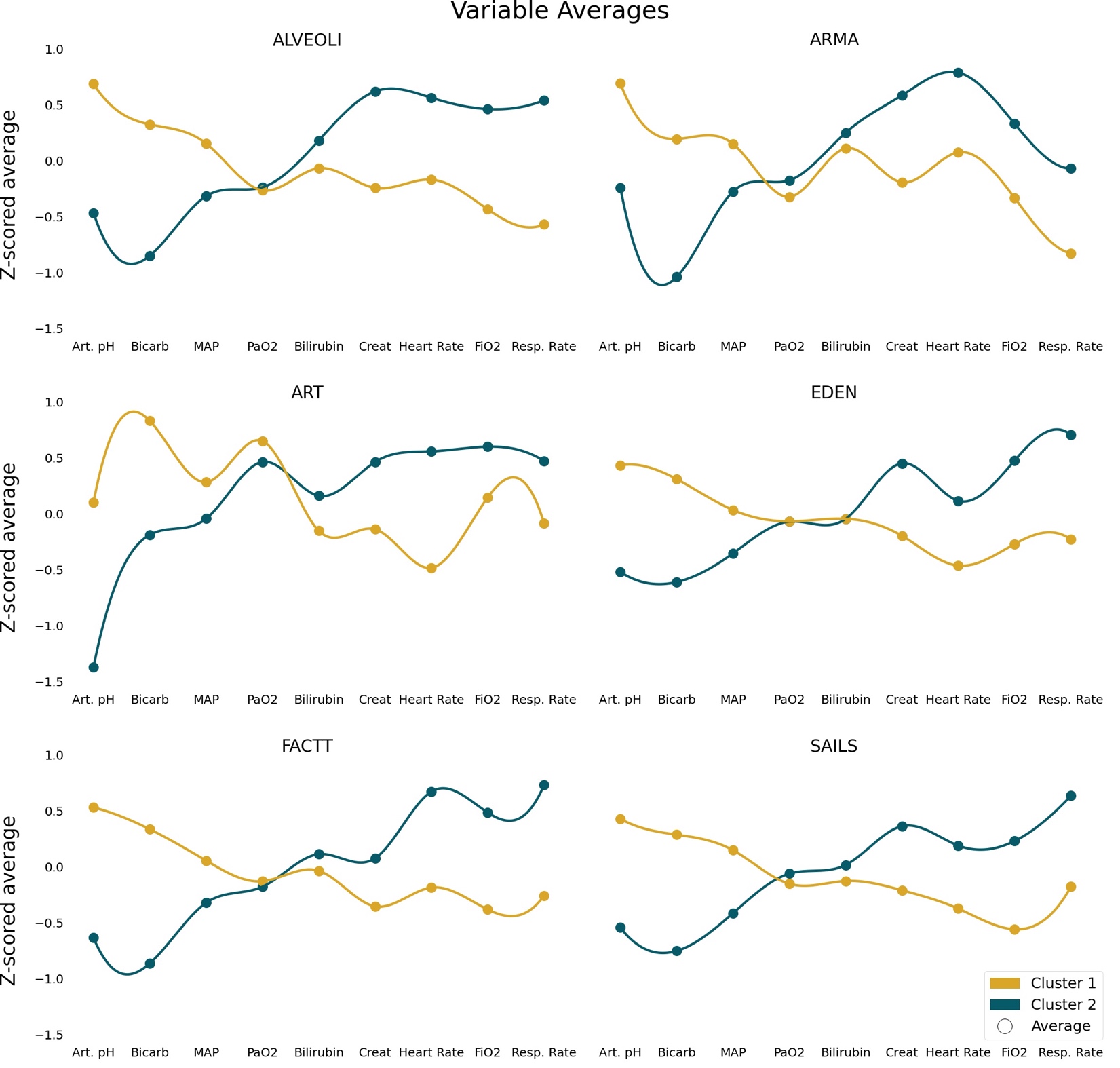

The circles represent the averages for each variable. The colored lines are exclusively to help visualize the opposite trends of the variables on the different clusters.

Abbreviations: Art. pH is arterial pH, Bicarb is bicarbonate, MAP is mean arterial pressure, Creat is creatinine and Resp. Rate is respiratory rate
